# Disaster and Disparities: The Disaster-Driven Inequality in Quality Antenatal Care Uptake in Bangladesh

**DOI:** 10.1101/2025.04.25.25326416

**Authors:** Md Injamul Haq Methun, Sanjida Akter, Ehsan Ahmed, Md Kamrul Hassan

## Abstract

**Introduction/Objectives:** This study assesses the impact of natural disasters on the inequality of quality ANC utilization.

**Methods:** This study uses data from the 2017-18 Bangladesh Demographic and Health Survey and monthly hazard reports from NIRAPAD. Inequalities in maternal healthcare utilization were measured using the Health Opportunity Index, and contributions of socioeconomic factors were decomposed using Shapley decomposition.

**Results:** The results reveal that inequality in the utilization of quality antenatal care is significantly greater in disaster-affected areas (32.72; 95% CI: 16.07, 49.38) compared to disaster-unaffected areas (27.17; 95% CI: 15.95, 38.39). Furthermore, socioeconomic factors such as place of residence, administrative division, maternal employment status, and birth order contribute more to these inequalities in disaster-affected regions than in unaffected areas.

**Conclusions:** The findings illustrate that natural disasters significantly increase the inequality in quality ANC utilization. Geographical location, employment status, and higher parity are key socio-economic factors contributing to the increasing inequality in quality ANC uptake. Therefore, Policymakers must prioritize equitable access to quality antenatal care by focusing on women of rural areas, those not engaged in formal employment, and individuals experiencing high-order pregnancies to mitigate these disaster-induced inequalities effectively.

## 1. Introduction

Natural disasters pose significant challenges to public health systems globally, particularly in low- and middle-income countries where resources and infrastructure are often limited. In Bangladesh, frequent natural disasters such as floods and cyclones disrupt essential healthcare services, exacerbating existing disparities and creating new challenges for vulnerable populations. Maternal healthcare, specifically antenatal care (ANC), is particularly susceptible to these disruptions, which can have severe consequences for both mothers and their children.

Bangladesh is highly vulnerable to natural disasters, with over 50 million people affected between 2000 and 2019 due to various catastrophic events. These disasters disrupt healthcare infrastructure and services, leading to significant declines in healthcare accessibility and quality. For instance, Brouwer et al. [1] noted increased health service inequality following earthquakes and floods, emphasizing the need for robust healthcare systems to withstand and recover from natural disasters.

The accessibility and quality of ANC are crucial determinants of maternal and neonatal health outcomes. Quality Antenatal Care (QANC) services play a critical role in ensuring a positive birth experience and mitigating adverse pregnancy outcomes such as anemia, preeclampsia, and low birth weight [2]. However, the disruptions caused by natural disasters often exacerbate existing healthcare disparities. For example, a study by Bhutta et al. [3] highlighted that in Ethiopia, post-disaster recovery of healthcare services was significantly hindered by pre-existing inequalities in healthcare access and quality.

Socioeconomic factors significantly influence healthcare access and utilization, and their impact is magnified in the wake of natural disasters. In disaster-affected areas, lower educational levels and poorer wealth status are more prevalent, contributing to reduced ANC utilization. For instance, the prevalence of primary education is lower in disaster-affected areas, and the proportion of the population in the poorest wealth quintile is higher. Ivers and Ryan [4] emphasized that socioeconomic vulnerabilities are often magnified post-disaster, further hindering access to essential healthcare services. This finding highlighting the need for targeted socio-economic interventions to mitigate these effects.

Rural residence is another critical factor contributing to disparities in QANC. Rural populations face significant challenges in accessing healthcare due to infrastructural damage and accessibility issues during and after disasters. Davis et al. [2] highlighted that rural healthcare infrastructure is often less resilient and more susceptible to disruption during disasters, necessitating targeted interventions to strengthen rural healthcare systems and ensure continuity of care. Employment opportunities are also crucial determinants of healthcare access post-disaster. Tran et al.[5] found that communities with higher levels of economic stability and employment were better able to recover and maintain access to healthcare services following disasters. This highlights the need for policies that support economic recovery and stability as a means of improving healthcare resilience.

Integrating maternal health services into disaster preparedness and response plans is vital for ensuring continuous and quality care during and after disasters. Comprehensive disaster risk reduction strategies that include maternal health components are essential. Zotti et al. [6] emphasized that disaster preparedness plans should include specific provisions for maternal and child health services to ensure these essential services are maintained even in the most challenging circumstances.

The critical need for integrating maternal health services into disaster preparedness and response plans cannot be overstated. Given the significant impact of natural disasters on healthcare access and quality, there is an urgent need to understand and address the disparities in ANC utilization in disaster-affected and unaffected areas of Bangladesh. This study aims to fill this gap by providing evidence-based insights that can inform policy and practice. By addressing the socioeconomic and infrastructural challenges exacerbated by disasters, policymakers and healthcare providers can develop targeted interventions to mitigate these disparities and improve health outcomes for vulnerable populations. Understanding these determinants at a regional level can help experts and organizations coordinate health-related activities and policies, ultimately leading to more effective and equitable healthcare systems in disaster-prone areas.

## 2. Methods

### 2.1 Data Source and Study Design

This study utilizes a cross-sectional design to analyze the effect of natural disasters on inequality in QANC utilization among reproductive-aged women of Bangladesh. The analysis was drowned on data from the Bangladesh Demographic and Health Survey 2017-18 (BDHS 2017-18), a nationally representative datasets that provide estimates of maternal and child health, nutrition, family planning, and fertility at the national and divisional levels. The study population included women aged 15 to 49 years who gave birth in the preceding three years of the survey. The data set does not contain any information that could identify individual participants during or after data collection. Besides the BDHS 2017-18 data, this study also utilizes the data on natural disasters from the monthly hazard reports of the Network for Information, Response, and Preparedness Activities on Disaster [7].

### 2.2 Sampling Procedure and Sample Size

The BDHS 2017-18 survey uses a two-stage stratified sampling procedure. Where urban and rural were the strata. In the first stage, 250 and 425 EAs were randomly selected from urban and rural strata. Each EA consists of, on average, 120 households. In each EA’s second stage, approximately 30 households were selected using systematic sampling. Thus, a total of 20,160 households were chosen for the interview, of which 19,457 households responded. Details of the sampling procedure are described elsewhere[8].

Among the selected households, 20,127 women were interviewed, of which 5052 were pregnant in the last three years before the survey. The study included 5,006 women, and the remaining 48 were excluded due to incomplete ANC information.

### 2.3 Variables

#### 2.3.1 Outcome Variable

Quality ANC is the outcome variable of this study. The QANC defined as BDHS 2017-18 is a woman who has at least four ANC visits with at least one medically trained provider and receives all essential components of ANC, which are weight and blood pressure measured, urine and blood sample taken, and informed about the signs of pregnancy complications [8]. The QANC is a binary variable. It takes a value of 1 if women received all of the above components; otherwise, it takes a value of 0.

#### 2.3.2 Exposure Variable

Disaster-affected is the exposure variable of this study. A List of upazila where the disasters occurred in the 3 years preceding the BDHS 2017-18 survey was made from the monthly hazard reports of NIRAPAD. Then, merging the GPS dataset of BDHS 2017-18 with the list of the variable disaster-affected created. Which takes value one if a woman experiences a natural disaster in this timeline; otherwise, it takes value 0.

#### 2.3.3 Covariates

The principle of equal opportunities states that everyone should have an equal chance to access the opportunities irrespective of their circumstances. Based on the literature[9– 14], in this study, socio-economic and demographic factors such as age, residence, division, education, sex of household head, wealth status, working status, husband’s education, family size, and birth order is considered as circumstances. Circumstances are the personal, family, or community characteristics one cannot directly affect [15].

### 2.4 Statistical Analysis

The inequality in quality ANC was assessed using the human opportunity index (HOI) and the dissimilarity index(D-index), which was developed by the World Bank in 2009 [15]. The inequality was decomposed using Shapley decomposition methods to determine the contributions of different circumstances on the inequality in the opportunity of QANC. The mathematical elaboration of the D-index, HOI-index, and Shapley decomposition was elsewhere [9]. Mann-Whitney U Test was used to identify the significant differences in HOI and D-index between disaster-affected and not-affected areas. Significance was considered at a p-value less than 0.05. The analysis was carried out in Stata version 17. We use QGIS 3.38 to merge the GIS data of BDHS 2017-18, and the shape file of BBS to identify the disaster-affected upazila.

## 3. Results

Background characteristics and reproductive status of women have presented in table 1. Most women reside in rural areas (65.6%). The 76.4% of women falls within the age group of 20–34 years. About 48% women have completed secondary education among them 46.9% were from disaster-affected areas. The majority of households (87.8%) have a male household head. About 62.5% women worked outside among them in disaster affected areas 61.5% women worked outside. Almost one-fifth of women were from poorest family. Approximately, one-third of husbands with primary and secondary education. In disaster-affected areas, 34.6% family have size 5-6 and 35.9% from family with more than 6 members. The highest percentage (56.9%) women pregnant with the second to fourth birth. Almost half of the(44.8%) women of disaster affected area reported that distance to healthcare facility was a big problem.

**Table 1.** Background characteristics and reproductive status of sample population.

| **Variables** | **Disaster-**<br>**unaffected Areas**<br>**n(%)** | **Disaster-affected**<br>**Areas**<br>**n(%)** | **Aggregate**<br>**n(%)** |
| --- | --- | --- | --- |
| **Age group** |  |  |  |
| 15-19 | 548(17.06) | 318(17.74) | 866(17.3) |
| 20-34 | 2475(77.03) | 1354(75.52) | 3829(76.49) |
| 35-49 | 190(5.91) | 121(6.75) | 311(6.21) |
| **Residence** |  |  |  |
| Urban | 1180(36.73) | 539(30.06) | 1719(34.34) |
| Rural | 2033(63.27) | 1254(69.94) | 3287(65.66) |
| **Division** |  |  |  |
| Barisal | 253(7.87) | 280(15.62) | 533(10.65) |
| Chattogram | 486(15.13) | 349(19.46) | 835(16.68) |
| Dhaka | 522(16.25) | 219(12.21) | 741(14.8) |
| Khulna | 391(12.17) | 133(7.42) | 524(10.47) |
| Mymensingh | 485(15.09) | 118(6.58) | 603(12.05) |
| Rajshahi | 382(11.89) | 139(7.75) | 521(10.41) |
| Rangpur | 315(9.8) | 244(13.61) | 559(11.17) |
| Sylhet | 379(11.8) | 311(17.35) | 690(13.78) |
| **Education** |  |  |  |
| No education | 188(5.85) | 124(6.92) | 312(6.23) |
| Primary education | 847(26.36) | 542(30.23) | 1389(27.75) |
| Secondary | 1559(48.52) | 841(46.9) | 2400(47.94) |
| Higher | 619(19.27) | 286(15.95) | 905(18.08) |
| **Sex of household head** |  |  |  |
| Male | 2831(88.11) | 1567(87.4) | 4398(87.85) |
| Female | 382(11.89) | 226(12.6) | 608(12.15) |
| **Wealth status** |  |  |  |
| Poorest | 576(17.93) | 503(28.05) | 1079(21.55) |
| Poorer | 650(20.23) | 365(20.36) | 1015(20.28) |
| Middle | 587(18.27) | 315(17.57) | 902(18.02) |
| Rich | 672(20.92) | 316(17.62) | 988(19.74) |
| Richest | 728(22.66) | 294(16.4) | 1022(20.42) |
| **Working Status** |  |  |  |
| Yes | 2027(63.09) | 1104(61.57) | 3131(62.54) |
| No | 1186(36.91) | 689(38.43) | 1875(37.46) |
| **Husband's highest education** |  |  |  |
| No education | 424(13.34) | 252(14.3) | 676(13.68) |
| Primary | 1031(32.44) | 623(35.36) | 1654(33.48) |
| Secondary | 1065(33.51) | 570(32.35) | 1635(33.1) |
| Higher | 648(20.39) | 314(17.82) | 962(19.47) |
| **Family Size** |  |  |  |
| 1-4 | 983(30.59) | 527(29.39) | 1510(30.16) |
| 5-6 | 1205(37.5) | 622(34.69) | 1827(36.5) |
| 6+ | 1025(31.9) | 644(35.92) | 1669(33.34) |
| **Parity/Birth Order** |  |  |  |
| 1 | 1231(38.31) | 680(37.93) | 1911(38.17) |
| 2-4 | 1836(57.14) | 1014(56.55) | 2850(56.93) |
| 5+ | 146(4.54) | 99(5.52) | 245(4.89) |
| **Getting medical help for self: distance to health facility** |  |  |  |
| Big problem | 1235(38.44) | 802(44.73) | 2037(40.69) |
| Not a big problem | 1978(61.56) | 991(55.27) | 2969(59.31) |

Table 2 present the association between socio-economic factors and quality Antenatal care. Utilization of QANC significantly differs by age group, place of residence, division respondents’ education, wealth status, working status, husband’s education, family size, birth order and distance to healthcare facility.

**Table 2.** Factors Affecting Quality ANC Utilization.

| **Variables** | **Received Quality ANC**<br>**n(%)** | **Did Not Receive Quality ANC**<br>**n(%)** | **P value** |
| --- | --- | --- | --- |
| **Age group** |  |  |  |
| 15-19 | 735(17.96) | 134(14.58) | <0.01 |
| 20-34 | 3092(75.54) | 740(80.52) |  |
| 35-49 | 266(6.5) | 45(4.9) |  |
| **Residence** |  |  |  |
| Urban | 1268(30.98) | 457(49.73) | <0.01 |
| Rural | 2825(69.02) | 462(50.27) |  |
| **Division** |  |  |  |
| Barisal | 449(10.97) | 84(9.14) | <0.01 |
| Chattogram | 698(17.05) | 137(14.91) |  |
| Dhaka | 562(13.73) | 179(19.48) |  |
| Khulna | 408(9.97) | 116(12.62) |  |
| Mymensingh | 497(12.14) | 106(11.53) |  |
| Rajshahi | 437(10.68) | 90(9.79) |  |
| Rangpur | 440(10.75) | 119(12.95) |  |
| Sylhet | 602(14.71) | 88(9.58) |  |
| **Education** |  |  |  |
| No education | 294(7.18) | 18(1.96) | <0.01 |
| Primary education | 1263(30.86) | 129(14.04) |  |
| Secondary | 1971(48.16) | 431(46.9) |  |
| Higher | 565(13.8) | 341(37.11) |  |
| **Sex of household head** |  |  |  |
| Male | 3608(88.15) | 796(86.62) | 0.19 |
| Female | 485(11.85) | 123(13.38) |  |
| **Wealth status** |  |  |  |
| Poorest | 1005(24.55) | 74(8.05) | <0.01 |
| Poorer | 918(22.43) | 99(10.77) |  |
| Middle | 760(18.57) | 145(15.78) |  |
| Rich | 770(18.81) | 218(23.72) |  |
| Richest | 640(15.64) | 383(41.68) |  |
| **Working Status** |  |  |  |
| Yes | 2520(61.57) | 612(66.59) | <0.01 |
| No | 1573(38.43) | 307(33.41) |  |
| **Husband's highest education** |  |  |  |
| No education | 628(15.58) | 51(5.57) | <0.01 |
| Primary | 1486(36.87) | 171(18.67) |  |
| Secondary | 1308(32.46) | 327(35.7) |  |
| Higher | 597(14.81) | 365(39.85) |  |
| **Family Size** |  |  |  |
| 1-4 | 1209(29.54) | 302(32.86) | 0.02 |
| 5-6 | 1530(37.38) | 300(32.64) |  |
| 6+ | 1354(33.08) | 317(34.49) |  |

| **Parity/Birth Order** |  |  |  |
| --- | --- | --- | --- |
| 1 | 1497(36.57) | 418(45.48) | <0.01 |
| 2-4 | 2364(57.76) | 488(53.1) |  |
| 5+ | 232(5.67) | 13(1.41) |  |
| **Getting medical help for self: distance to health facility** |  |  |  |
| Big problem | 1769(43.22) | 271(29.49) | <0.01 |
| Not a big problem | 2324(56.78) | 648(70.51) |  |
| **Disaster Affected Area** |  |  |  |
| Yes | 2582(63.18) | 631(68.66) | <0.01 |
| No | 1505(36.82) | 288(31.34) |  |

Disaster affected areas showed a significant association with QANC. In Disaster affected areas, women received highest percentage (63.1%) of QANC and 36.8% women from non-disaster affected areas received QANC.

Natural disasters have significant effect on the access to the QANC and equality in the utilization of the QANC (Table 3). Women of disaster-unaffected area (19.82; 95% CI: 18.52, 21.13) has significant higher access than the women located in disaster-affected area (16.23; 95% CI: 14.62, 17.85). Disasters exacerbate the inequality in QANC uptake, as the dissimilarity index of disaster-affected area (32.72; 95% CI: 16.07, 49.38)) was higher than the disaster unaffected area (27.17; 95% CI: 15.95, 38.39).

**Table 3:**
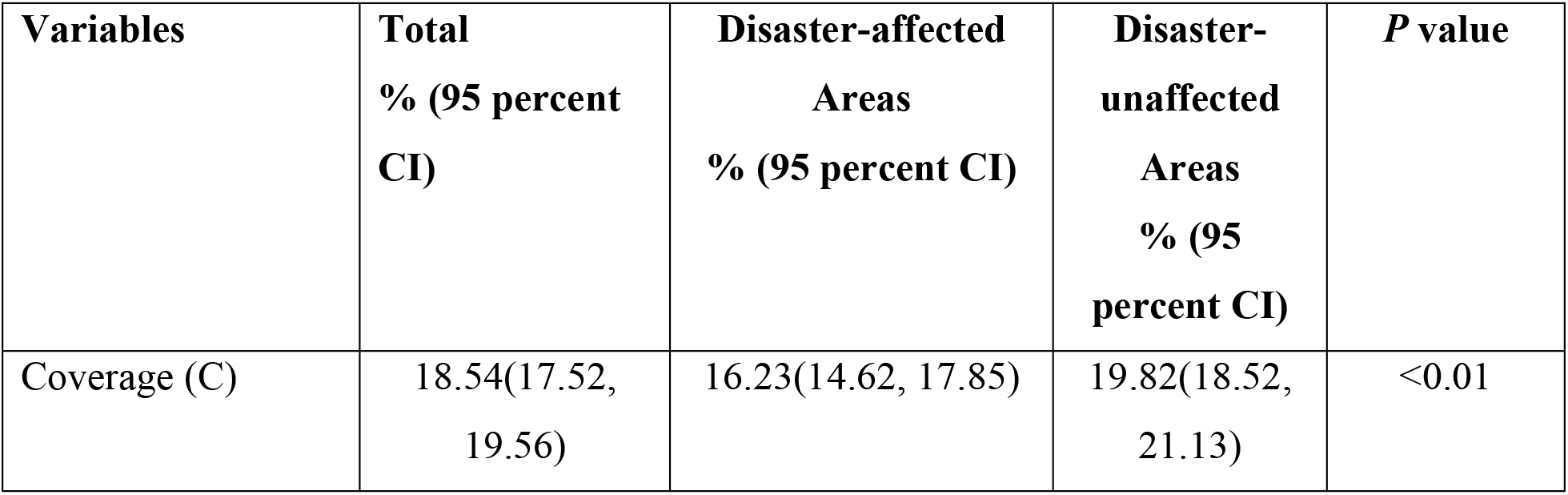
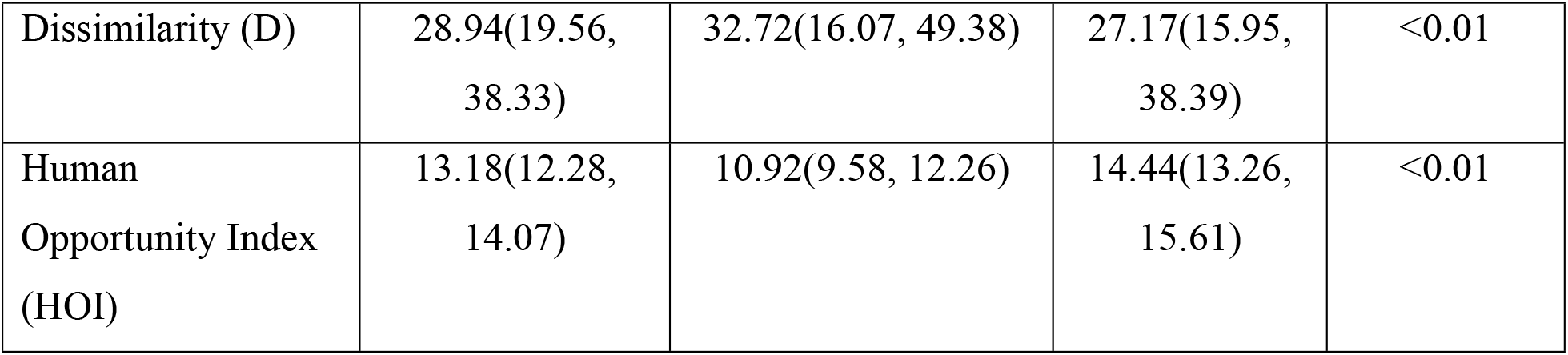
Disparities in Quality ANC among Disaster-affected and unaffected populations.

| **Variables** | **Total**<br>**% (95 percent**<br>**CI)** | **Disaster-affected**<br>**Areas**<br>**% (95 percent CI)** | **Disaster-**<br>**unaffected**<br>**Areas**<br>**% (95**<br>**percent CI)** | ***P* value** |
| --- | --- | --- | --- | --- |
| Coverage (C) | 18.54(17.52,<br>19.56) | 16.23(14.62, 17.85) | 19.82(18.52,<br>21.13) | <0.01 |
| Dissimilarity (D) | 28.94(19.56, 38.33) | 32.72(16.07, 49.38) | 27.17(15.95, 38.39) | <0.01 |
| Human Opportunity Index (HOI) | 13.18(12.28, 14.07) | 10.92(9.58, 12.26) | 14.44(13.26, 15.61) | <0.01 |

Table 4 displays the impact of disasters on the socio-economic factors’ contributions to inequality of the QANC utilization. Among the socio-economic factors contributing to the inequalities in the QANC utilization, the contributions of residence (4.31; 95% CI: 2.98, 5.64), Division (2.84; 95% CI: 1.99, 3.69), wealth status (2.8; 95% CI: 0.10, 5.52), working status (8.07; 95% CI: 6.69, 9.45) and birth order (3.16, 95% CI: 1.63, 4.69) were higher in disaster affected areas compared to the disaster unaffected-areas. Whereas, the contributions of husband’s education (−3.74; 95% CI: −5.99, −1.49) and problems due to distance to health facility (−5.51; 95% CI:-6.89, −4.1) were reduced in disaster affected area than the disaster unaffected area.

**Table 4:** Comparison of Socio-Demographic Factors’ Contributions to the Inequality of Quality ANC between Disaster-Affected and Unaffected Areas.

| Variables | Disaster-Affected Area | Disaster-Unaffected Area | Difference | P value |
| --- | --- | --- | --- | --- |
| Age group | 0.7 | 0.96 | 0.26(-0.30, 0.82) | 0.21 |
| Residence | 6.84 | 2.53 | 4.31 (2.98, 5.64) | <0.00 |
| Division | 3.05 | 0.21 | 2.84(1.99, 3.69) | <0.00 |
| Education | 19.55 | 20.37 | 0.82 (-1.53, 3.17) | 0.26 |
| Sex of household | 0.22 | 0.84 | 0.62 (0.19, 1.05) | <0.00 |
| Wealth status | 31.81 | 29.00 | 2.81 (0.10, 5.52) | 0.02 |
| Working Status | 8.76 | 0.69 | 8.07 (6.69, 9.45) | <0.00 |
| Husband's highest education | 16.48 | 20.22 | 3.74 (1.49, 5.99) | <0.00 |
| Family Size | 0.51 | 0.74 | 0.23 (-0.26, 0.72) | 0.22 |
| Getting Medical help for self: distance to health facility | 3.79 | 9.3 | 5.51 (4.13, 6.89) | <0.00 |
| Birth order | 8.3 | 5.14 | 3.16 (1.63, 4.69) | <0.00 |

## 4. Discussion

The study reveals several key findings: lower coverage of quality antenatal care (ANC) in disaster-affected areas compared to unaffected areas, higher Dissimilarity Index in disaster-affected areas compared to unaffected areas, significant impact of working status on QANC utilization in disaster-affected areas, and a more substantial impact of residence, division, lower educational levels, and wealth status on ANC inequality in disaster-affected areas.

The study shows lower coverage of QANC in disaster-affected areas compared to unaffected areas. This finding is in line with Carballo et al.[16], who demonstrated that natural disasters significantly disrupt healthcare services which leads to reduced access to essential care services during critical periods.

The higher Dissimilarity Index in disaster-affected areas compared to unaffected areas in this study indicates greater inequality in access to QANC. Similarly, HOI is lower in disaster-affected areas compared to unaffected areas. This is consistent with the work of Rocklöv et al. [17], who found that natural disasters often exacerbate existing health disparities, leading to increased inequality in health service utilization.

The comparative analysis reveals that residence, division, and wealth status have a more substantial impact on ANC inequality in disaster-affected areas. This finding is supported by Phalkey et al. [18], who highlighted that rural populations and those in lower socio-economic brackets are disproportionately affected by natural disasters, resulting in greater healthcare disparities. Phalkey et al. argued that rural healthcare infrastructure is often less resilient and more susceptible to disruption, exacerbating disparities in healthcare access during disasters. Their research suggests that strengthening rural healthcare systems should be a priority in disaster risk reduction strategies.

Research by Shoaf and Rottman[19] further corroborates these findings, demonstrating that access to healthcare services diminishes significantly in the wake of natural disasters, with rural and economically disadvantaged populations being the most affected. This is particularly relevant in the context of this study, where rural residence and lower socio-economic status are prevalent in disaster-affected areas, contributing to the observed disparities in QANC.

In the context of maternal health, the findings of lower QANC coverage and higher disparities in disaster-affected regions align with those of Bobo et al. [20], who found that natural disasters lead to increased maternal morbidity and mortality, primarily due to disrupted healthcare services and reduced access to quality care. Similarly, Chand & Loosemore [21], highlighted that the lack of resilient healthcare infrastructure exacerbates these issues, making it challenging to provide continuous care during and after disasters.

The findings indicate a significant impact of working status on QANC utilization in disaster-affected areas, align with those of Bangalore et al. [22]. They emphasized that economic resilience and recovery significantly influence the ability of populations to access healthcare services after a disaster. They argued that policies aimed at economic recovery should include measures to support healthcare systems, ensuring that economic growth translates into improved health outcomes.

The findings lower educational levels in disaster-affected areas contribute to lower QANC utilization supports the assertions of Cutter et al. [23], who found that higher levels of education are associated with better health outcomes and increased healthcare utilization, even in disaster-affected areas. They highlighted the role of education in enhancing health literacy, which in turn improves individuals’ ability to seek and utilize healthcare services effectively.

Finally, this study highlights the critical need for integrating maternal health services into disaster preparedness and response plans. This is supported by research by Salama et al. [24] who emphasized that comprehensive disaster risk reduction strategies that include maternal health components are essential for ensuring continuous and quality care during and after disasters.

## 5. Conclusion

This study assessed the impact of natural disaster on the inequality of QANC utilization in Bangladesh using nationally representative 2017-18 Bangladesh Demographic and Health Survey 2017-18 data. The findings demonstrate that there is significant inequalities in QANC uptake in Bangladesh. Women reside in disaster affected areas faced higher inequality in QANC uptake compared to the inequality faced by women in disaster unaffected areas. The influence of geographic location, working status and parity on QANC uptake inequality significantly increase in disaster affected area. Whereas, the contributions of education and distance to the health facilities were significantly decrease in disaster affected areas compared to the unaffected areas.

The results highlight that the policy makers need to come up with targeted policy intervention to address the vulnerabilities of women in rural areas, those not engaged in formal employment, and individuals experiencing high-order pregnancies. Since, these women are disproportionately affected by the adverse impacts of natural disasters on quality maternal healthcare utilization. To ensure the equitable access to quality antenatal care and resilience in maternal health services it is important to strengthen the healthcare system in vulnerable areas especially in disaster-affected rural areas.

Despite the well-designed methodology, this study has some limitations. This study cannot find the causal effect of disaster since it uses cross-sectional data. This study finds out short term impact of disaster on QANC uptake inequalities. Further research may explore long-term impact disaster on the inequality of maternal health care utilization using longitudinal study. Overall, this study highlights the critical need to give importance on rural disaster affected area into maternal health policies to achieve equitable healthcare outcomes in Bangladesh.

## 6. Ethics approval and consent to participate

Not applicable.

## 7. Consent for publication

Not applicable for this study.

## 8. Availability of data

The datasets analyzed during the current study are available in the Demographic and Health Surveys (DHS) Program repository https://dhsprogram.com/data/. DHS data are publicly available upon request.

## 10. Author Contributions

**Md Injamul Haq Methun:** Conceptualization, Data curation, Formal analysis, Methodology, Writing – original draft, Writing – review & editing.

**Sanjida Akter:** Data curation, Writing – original draft.

**Ehsan Ahmed:** Formal analysis, Writing – original draft.

**Md Kamrul Hassan:** Formal analysis, Writing – original draft, Writing – review & editing.

## 11. Acknowledgments

The authors want to acknowledge the Measures DHS data archive for providing us with the datasets for further analysis.

## References

1. Brouwer R, Akter S, Brander L, Haque E. Socioeconomic vulnerability and adaptation to environmental risk: a case study of climate change and flooding in Bangladesh. Risk Anal. 2007;27: 313–326. doi:10.1111/J.1539-6924.2007.00884.X

2. Davis JR, Wilson S, Brock-Martin A, Glover S, Svendsen ER. The Impact of Disasters on Populations With Health and Health Care Disparities. Disaster Med Public Health Prep. 2010;4: 30–38. doi:10.1017/S1935789300002391

3. Bhutta ZA, Das JK, Rizvi A, Gaffey MF, Walker N, Horton S, et al. Evidence-based interventions for improvement of maternal and child nutrition: what can be done and at what cost? Lancet. 2013;382: 452–477. doi:10.1016/S0140-6736(13)60996-4

4. Ivers LC, Ryan ET. Infectious diseases of severe weather-related and flood-related natural disasters. Curr Opin Infect Dis. 2006;19: 408–414. doi:10.1097/01.QCO.0000244044.85393.9E

5. Tran P, Marincioni F, Shaw R, Sarti M, An L Van. Flood risk management in Central Viet Nam: Challenges and potentials. Natural Hazards. 2008;46: 119–138. doi:10.1007/S11069-007-9186-2/METRICS

6. Zotti ME, Williams AM, Wako E. Post-disaster Health Indicators for Pregnant and Postpartum Women and Infants. Matern Child Health J. 2015;19: 1179–1188. doi:10.1007/S10995-014-1643-4/METRICS

7. NIRAPAD. Available: https://www.nirapad.org.bd/home/resources/monthlyHazard

8. of Population Research NI, NIPORT T-, of Health M, Welfare F, ICF. Bangladesh Demographic and Health Survey 2017-18. 2020. Available: https://dhsprogram.com/publications/publication-FR344-DHS-Final-Reports.cfm

9. Methun MIH, Ahinkorah BO, Roy S, Okyere J, Hossain MI, Haq I, et al. Inequalities in adequate maternal healthcare opportunities: evidence from Bangladesh Demographic and Health Survey 2017–2018. BMJ Open. 2023;13: e070111. doi:10.1136/BMJOPEN-2022-070111

10. Katemba BM, Bwembya P, Hamoonga TE, Chola M, Jacobs C. Demand Side Factors Associated With Quality Antenatal Care Services: A Case Study of Lusaka District, Zambia. Front Public Health. 2018;6: 361081. doi:10.3389/FPUBH.2018.00285/BIBTEX

11. Negash WD, Fetene SM, Shewarega ES, Fentie EA, Asmamaw DB, Teklu RE, et al. Multilevel analysis of quality of antenatal care and associated factors among pregnant women in Ethiopia: a community based cross-sectional study. BMJ Open. 2022;12: e063426. doi:10.1136/BMJOPEN-2022-063426

12. Joshi C, Torvaldsen S, Hodgson R, Hayen A. Factors associated with the use and quality of antenatal care in Nepal: A population-based study using the demographic and health survey data. BMC Pregnancy Childbirth. 2014;14: 1–11. doi:10.1186/1471-2393-14-94/TABLES/3

13. Simkhada B, Teijlingen ER Van, Porter M, Simkhada P. Factors affecting the utilization of antenatal care in developing countries: systematic review of the literature. J Adv Nurs. 2008;61: 244–260. doi:10.1111/J.1365-2648.2007.04532.X

14. Raru TB, Ayana GM, Bahiru N, Deressa A, Alemu A, Birhanu A, et al. Quality of antenatal care and associated factors among pregnant women in East Africa using Demographic and Health Surveys: A multilevel analysis. Womens Health (Lond). 2022;18. doi:10.1177/17455065221076731

15. Vega JRM, Barros RP De, Chanduvi JS, Cord LJ. Do our children have a chance?: A human opportunity report for Latin America and the Caribbean. 2011. Available: https://books.google.com/books?hl=en&lr=&id=uZH4XTNyvsIC&oi=fnd&pg=PR5&dq=10.1596/978-0-8213-8699-6&ots=9TVubdmGrB&sig=t3Tch0t5MAw_Z4DQxnjqNXzmEIM

16. Carballo M, Daita S, Hernandez M, Soc JR. Impact of the Tsunami on Healthcare Systems. https://doi.org/101177/014107680509800902. 2005;98: p390–395. doi:10.1177/014107680509800902

17. Rocklöv J, Sauerborn R, Sankoh O. Guest Editorial: Weather conditions and population level mortality in resource-poor settings – understanding the past before projecting the future. Glob Health Action. 2012;5: 2–5. doi:10.3402/GHA.V5I0.20010

18. Phalkey RK, Aranda-Jan C, Marx S, Höfle B, Sauerborn R. Systematic review of current efforts to quantify the impacts of climate change on undernutrition. Proc Natl Acad Sci U S A. 2015;112: E4522–E4529. doi:10.1073/PNAS.1409769112/SUPPL_FILE/PNAS.1409769112.ST02.DOCX

19. Shoaf KI, Rottman SJ. The Role of Public Health in Disaster Preparedness, Mitigation, Response, and Recovery. Prehosp Disaster Med. 2000;15: 18–20. doi:10.1017/S1049023X00025243

20. Bobo FT, Asante A, Woldie M, Hayen A. Poor coverage and quality for poor women: Inequalities in quality antenatal care in nine East African countries. Health Policy Plan. 2021;36: 662–672. doi:10.1093/HEAPOL/CZAA192

21. Chand AM, Loosemore M. Hospital disaster management’s understanding of built environment impacts on healthcare services during extreme weather events. Engineering, Construction and Architectural Management. 2016;23: 385–402. doi:10.1108/ECAM-05-2015-0082/FULL/XML

22. Bangalore M, Hallegatte S, Vogt-Schilb A, Rozenberg J. Unbreakable: Building the Resilience of the Poor in the Face of Natural Disasters. Unbreakable: Building the Resilience of the Poor in the Face of Natural Disasters. 2017. doi:10.1596/978-1-4648-1003-9

23. Cutter SL, Boruff BJ, Shirley WL. Social vulnerability to environmental hazards. Soc Sci Q. 2003;84: 242–261. doi:10.1111/1540-6237.8402002

24. Salama P, Spiegel P, Talley L, Waldman R. Lessons learned from complex emergencies over past decade. Lancet. 2004;364: 1801–1813. doi:10.1016/S0140-6736(04)17405-9

